# Association of Obstructive Sleep Apnea and severity of COVID-19: A hospital based observational study

**DOI:** 10.1101/2020.11.12.20230631

**Authors:** Avishek Kar, Khushboo Saxena, Abhishek Goyal, Abhijit Pakhare, Alkesh Khurana, Saurabh Saigal

**Affiliations:** Department of Pulmonary Medicine, AIIMS Bhopal; Pulmonary Medicine, AIIMS Bhopal; CFM, AIIMS Bhopal; Anesthesiology, AIIMS Bhopal

**Keywords:** COVID-19, STOP BANG, NoSAS, ESS, OSA, Berlin Questionnaire

## Abstract

**Introduction:** OSA has been postulated to be associated with mortality in COVID19, but studies are lacking thereof. This study was done to estimate prevalence of OSA in patients with COVID-19 using various screening questionnaires and to assess effect of OSA on outcome of disease.

**Methodology:** In this prospective observational study, consecutive patients with RTPCR confirmed COVID 19 patients were screened for OSA by different questionnaires (STOPBANG, Berlin Questionnaire, NoSAS and Epworth Scale). Association between OSA and outcome (mortality) and requirement for respiratory support was assessed.

**Results:** In study of 213 patients; screening questionnaires for OSA {STOPBANG, Berlin Questionnaire (BQ), NoSAS} were more likely to be positive in patients who died compared to patients who survived. On binary logistic yregression analysis, age≥55 and STOPBANG score ≥5 were found to have small positive but independent effect on mortality even after adjusting for other variables. Proportion of patients who were classified as high risk for OSA by various OSA screening tools significantly increased with increasing respiratory support (p<0.001 for STOPBANG, BQ, ESS and p=0.004 for NoSAS).

**Conclusion:** This is one of the first prospective studies of sequentially hospitalized patients with confirmed COVID 19 status who were screened for possible OSA. This study shows that OSA could be an independent risk factor for poor outcome in patients with COVID19.

## Introduction

The beginning of 2020 saw the evolution of COVID-19 into a global pandemic. Severe acute respiratory syndrome coronavirus 2(SARS-CoV-2) was discovered in the Chinese province of Wuhan in December 2019 and is responsible for the causation of coronavirus disease 2019(COVID-19)^1^. As of October 24,2020, over 42 million people spread across 215 countries around the world have been affected by COVID-19 resulting in 1,141,567 deaths.^2^ As the world struggles to cope with this pandemic, researchers across the world are curetting mechanistic pathways that would possibly explain the disease severity in certain population.

Mortality in Covid-19 disease was mainly seen in the subgroup of patients who developed severe respiratory failure owing to acute interstitial pneumonia involving both lungs and acute respiratory distress syndrome(ARDS)^3^.While the understanding of pathogenesis is still a topic of research, few studies have pointed clinical association between mortality and older age, male sex, hypertension, diabetes, obesity and cardiovascular diseases.^4,5,6,7^InterestinglyObstructive Sleep Apnea (OSA)is also commonly associated with these similar comorbidities.

OSA is a pro-inflammatory state involving mediators like IL-6, TNF-alpha, MCP-1 etc^8^. IL-6 levels have also got prognostic implications in COVID-19 disease indicating that OSA might lead to worsening hypoxemia and cytokine storm. Furthermore, it has been hypothesized that SARS-CoV-2 infects humans through ACE-2 receptor which again has increased expression in obese OSA patients^9,10^. Obesity causes impaired respiratory mechanics leading to decreased FEV1, FVC, diaphragmatic excursion which in turn worsens outcome in patients with respiratory failure^11^. OSA can cause hypoxemia leading to poorer outcome in patients of COVID-19 pneumonia.

In accordance with these facts, in a recently concluded study it was observed that almost one third of the COVID-19 cases requiring Intensive Care Unit (ICU) admission had pre-existent OSA^12^. Similarly in two small studies on patients of severe COVID-19 pneumonia it was observed that one quarter of the patients were known case of OSA^13,14^.

Since OSA and patients developing COVID19 ARDS have so much in common, it is worthwhile to look into the association between these two. Thus, we need prospective studies to see whether people with OSA more at risk are to develop COVID19 related complications. If any association is found, this will further help in early triaging of complication prone population and possibly in prevention of complications. This study was planned to estimate proportion of COVID-19 patients who have OSA based on various standard screening questionnaires and to explore if there is any association of being at high risk for OSA and severity of COVID-19 including mortality.

## Methods

### Setting and design

This was a single center prospective observational study done at All India Institute of Medical Sciences Bhopal, India between 10^th^ August and 22^nd^ September 2020 on consecutive COVID 19 positive patients admitting to intensive care as well as isolation wards of hospital.

### Participants and procedures

All consecutive patients with confirmed positive report of COVID 19 RT-PCR on nasopharyngeal and oropharyngeal swab were enrolled. Patients were either admitted to intensive care or isolation wards as per clinical decision. Inclusion and exclusion criteria were as follows

#### Inclusion criteria

1. Patients positive for COVID 19 by RT-PCR of nasopharyngeal and oropharyngeal swab.
2. Age >18 years.
3. Patients whose spouse or bed partner was willing for confirmation of history and sleeping pattern.
4. Patients and attendants who gave written informed consent for participating in study.

#### Exclusion Criteria

1. Patients/ attendants who refused to cooperate or to give written informed consent
2. Sleeping partner not available to confirm history given by patient
3. Patient who were already intubated and were on mechanical ventilation
4. Patients who were not in a state to answer questions
5. Any recent surgery in last one month

After obtaining informed consent from patient or bed partner of patient, demographic details, medical history including co-morbidities were noted at the time of admission. As general practice, height and weight is measured for all patients in our ICU and ward at the time of admission. BMI was calculated according to other recorded weight and height data. Height and weight were measured using Seca^®^213 portable stadiometer and Seca^®^803 electronic flat scale respectively. Neck circumference was measured at the level of cricothyroid membrane in sitting position using girth measuring tape.

Patients were asked for NOSAS^15^, STOP BANG^16^, BERLIN^17^ and Epworth sleepiness scores^18^ within one day of admission. History given by patient was reconfirmed by bed partner of same patient.

NoSAS score category was classified as OSA if score was ≥8. STOP BANG was classified further as high risk for score ≥ 5. Berlin score was calculated in all three categories and classified into high risk if there were two or more categories with score ≥ 2 and low risk if there was only one category or no category where score was ≥ 2. ESS scores≥ 10 was used to classify ESS into high or low risk categories.

Patients were observed for their maximum oxygen and ventilatory requirement during their stay in hospital and were further divided into four groups i.e.

1. ‘No Oxygen’ group: Patients who did not require oxygen during their hospital stay.
2. ‘Oxygen Only’ group: Patients who required oxygen through facemask, venture mask or nasal prongs and their fraction of oxygen requirement was less than 0.5. These Patients never required Invasive Mechanical Ventilation (IMV) or Non-Invasive Ventilation (NIV) or High Flow Nasal Cannula (HFNC) during their hospital stay.
3. ‘NIV’ group: Patients who required either NIV or HFNC anytime during their stay. These patients were never intubated during their hospital stay.
4. ‘IMV’ group: Patients who were intubated.

Patients were followed up until their final outcome in the form of either discharge from hospital or death.

### Data analysis

We have used R software version 3.6.1 with *gtsummary, ggplot2* and *finalfit* packages for data analysis^19,20,21,22^. Nominal variables are summarized as count and percentages while numerical variables as mean and standard deviation. Proportion of patients with high risk for OSA as defined by different questionnaires were analyzed and compared for baseline characteristics, types of respiratory or ventilatory requirement and mortality. Difference in distribution of nominal variables across groups was tested by Chi-square test and in numerical variables by Wilcoxan rank sum test. Logistic regression models were fitted separately to estimate effect age, gender, neck circumference, history of diabetes, hypertension, coronary artery disease and STOP BANG score categories. Then to multivariable logistic regression model was fitted to test effect of variables which had p<0.25 in univariable analysis. Model assumptions and goodness-of-fit was also tested by standard procedures.

### Ethics and permissions

Institutional Human Ethics Committee of AIIMS Bhopal reviewed and approved study protocol with approval letter number IHEC-LOP/2020/IM0309. Eligible patients and attendants were provided participant information sheet in native language for explaining purpose of study, procedures and expectations from participants.

## Results

During study period, 250 patients with RT PCR positive for COVID19 got admitted in our hospital. After applying inclusion and exclusion criteria, 213 patients (144 male and 69 female) were finally enrolled and were prospectively followed till final outcome (discharge/death). Out of 213 patients, 57 succumbed due to COVID19 ARDS (Table1). Patients who died were elderly (p<0.001) and were more likely to have hypertension (p=0.007) and/or Diabetes mellitus (p=0.001). Screening questionnaires for OSA {STOPBANG, Berlin Questionnaire (BQ), NoSAS} were more likely to be positive in patients who died compared to patients who survived. Similarly, Epworth sleepiness scale (ESS) score was significantly higher in deceased group {12(9.0-13.0) v/s 9.0(5.2-11.0)} (p<0.001).

**Table 1:** Baseline characteristics (Demographic, OSA screening tool scores and clinical) among survived and deceased patients

| Characteristic | Overall<br>(N = 213) | Survived<br>(n= 156) | Deceased<br>(n = 57) | p-value |
| --- | --- | --- | --- | --- |
| Age | 55 (44, 64) | 53 (40, 62) | 60 (52, 69) | <0.001 |
| Gender |  |  |  | >0.9 |
| Male | 144 (68%) | 105 (67%) | 39 (68%) |  |
| Female | 69 (32%) | 51 (33%) | 18 (32%) |  |
| Diabetes | 93 (44%) | 59 (38%) | 34 (60%) | 0.007 |
| Hypertension | 94 (44%) | 58 (37%) | 36 (63%) | 0.001 |
| CAD | 25 (12%) | 14 (9.0%) | 11 (19%) | 0.067 |
| CKD | 5 (2.3%) | 1 (0.6%) | 4 (7.0%) | 0.019 |
| BMI | 26.7 (23.6, 29.5) | 26.3 (23.6, 29.4) | 28.0 (24.1, 30.1) | 0.2 |
| Neck Circumference | 40.0 (38.0, 42.0) | 40.0 (37.0, 42.0) | 41.0 (38.0, 43.0) | 0.007 |
| STOP BANG | 3.00 (2.00, 4.00) | 3.00 (2.00, 4.00) | 4.00 (3.00, 5.00) | <0.001 |
| STOP BANG<br>Category |  |  |  | <0.001 |
| Low to Intermediate | 160 (77%) | 129 (84%) | 31 (55%) |  |
| High | 49 (23%) | 24 (16%) | 25 (45%) |  |
| Berlin Category |  |  |  | 0.002 |
| Low Risk | 108 (51%) | 90 (58%) | 18 (33%) |  |
| High Risk | 102 (49%) | 65 (42%) | 37 (67%) |  |
| NoSAS | 9.0 (6.0, 13.0) | 9.0 (6.0, 11.0) | 11.0 (8.0, 15.0) | <0.001 |
| NoSAS Category |  |  |  | 0.020 |
| No OSA | 69 (33%) | 58 (38%) | 11 (20%) |  |
| OSA | 140 (67%) | 95 (62%) | 45 (80%) |  |
| ESS Score | 9.0 (6.0, 12.0) | 9.0 (5.2, 11.0) | 12.0 (9.0, 13.0) | <0.001 |
| ESS Category |  |  |  | <0.001 |
| High Risk | 98 (47%) | 60 (39%) | 38 (68%) |  |
| Low Risk | 112 (53%) | 94 (61%) | 18 (32%) |  |

Similar findings were seen when baseline characteristics of 213 patients were compared according to across various modes of respiratory support (without oxygen, oxygen only, NIV group or IMV groups) (Table2).

**Table 2:** Baseline characteristics (Demographic, OSA screening tool scores and clinical) across various modes of respiratory support

| Characteristic | No Oxygen,<br>N = 71 | Oxygen,<br>N = 43 | NIV,<br>N = 37 | IMV,<br>N = 62 | p-value |
| --- | --- | --- | --- | --- | --- |
| Age | 46 (32, 62) | 57 (50, 66) | 53 (50, 60) | 59 (51, 67) | <0.001 |
| Male | 48 (68%) | 30 (70%) | 23 (62%) | 43 (69%) | 0.9 |
| Female | 23 (32%) | 13 (30%) | 14 (38%) | 19 (31%) |  |
| Diabetes | 11 (15%) | 24 (56%) | 22 (59%) | 36 (58%) | <0.001 |
| Hypertension | 13 (18%) | 22 (51%) | 22 (59%) | 37 (60%) | <0.001 |
| CAD | 4 (5.6%) | 6 (14%) | 5 (14%) | 10 (16%) | 0.2 |
| CKD | 0 (0%) | 0 (0%) | 1 (2.7%) | 4 (6.5%) | 0.035 |
| Neck Circumference | 39.0 (37.0, 41.0) | 39.5 (37.0, 42.0) | 40.0 (39.0, 42.0) | 40.0 (38.0, 43.0) | 0.067 |
| STOP BANG Score | 2.00 (1.00, 3.00) | 4.00 (2.00, 4.00) | 4.00 (3.00, 4.00) | 4.00 (3.00, 5.00) | <0.001 |
| STOP BANG Group |  |  |  |  |  |
| Low to Intermediate | 65 (92%) | 33 (79%) | 27 (77%) | 35 (57%) | <0.001 |
| High | 6 (8.5%) | 9 (21%) | 8 (23%) | 26 (43%) |  |
| Berlin Category |  |  |  |  |  |
| Low Risk | 57 (80%) | 18 (42%) | 12 (33%) | 21 (35%) | <0.001 |
| High Risk | 14 (20%) | 25 (58%) | 24 (67%) | 39 (65%) |  |
| NoSAS | 8.0 (3.5, 11.0) | 9.0 (7.0, 13.0) | 11.0 (7.5, 12.5) | 11.0 (8.0, 14.0) | <0.001 |
| NoSAS Category |  |  |  |  |  |
| No OSA | 35 (49%) | 12 (29%) | 9 (26%) | 13 (21%) | 0.004 |
| OSA | 36 (51%) | 30 (71%) | 26 (74%) | 48 (79%) |  |
| ESS Score | 6.0 (4.0, 10.0) | 9.0 (8.0, 11.0) | 9.5 (8.0, 12.2) | 12.0 (9.0, 13.0) | <0.001 |
| High ESS (>10) | 20 (28%) | 20 (48%) | 18 (50%) | 40 (66%) | <0.001 |
| Low ESS (<10) | 51 (72%) | 22 (52%) | 18 (50%) | 21 (34%) |  |

Proportion of patients who were classified as high risk for OSA by various OSA screening tools significantly increased with increasing respiratory support (Table 2 and Figure 2) (p<0.001 for STOPBANG, BQ, ESS and p=0.004 for NoSAS).

**Figure 1:**
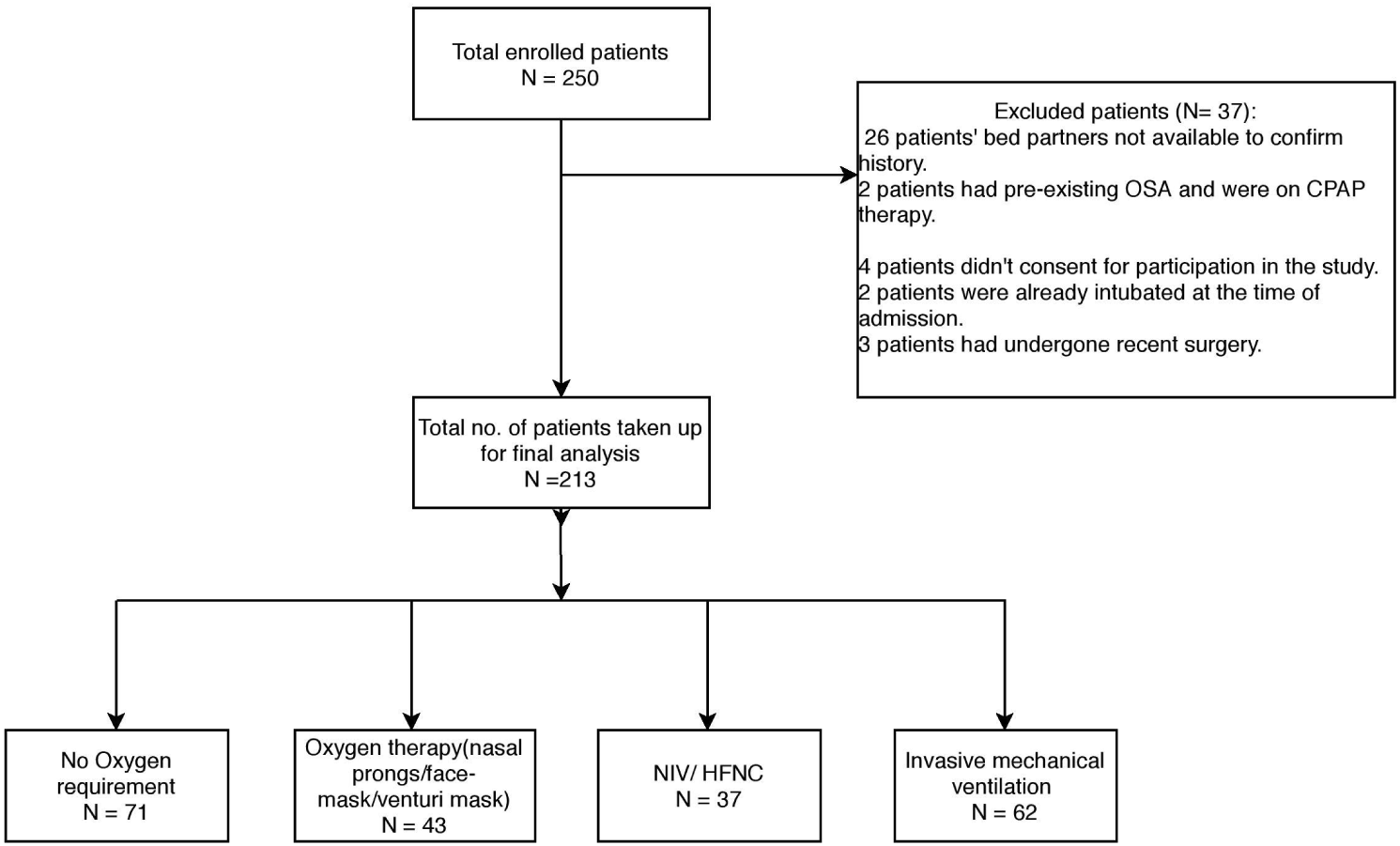
Flowchart of patients admitted during study period.

**Figure 2:**
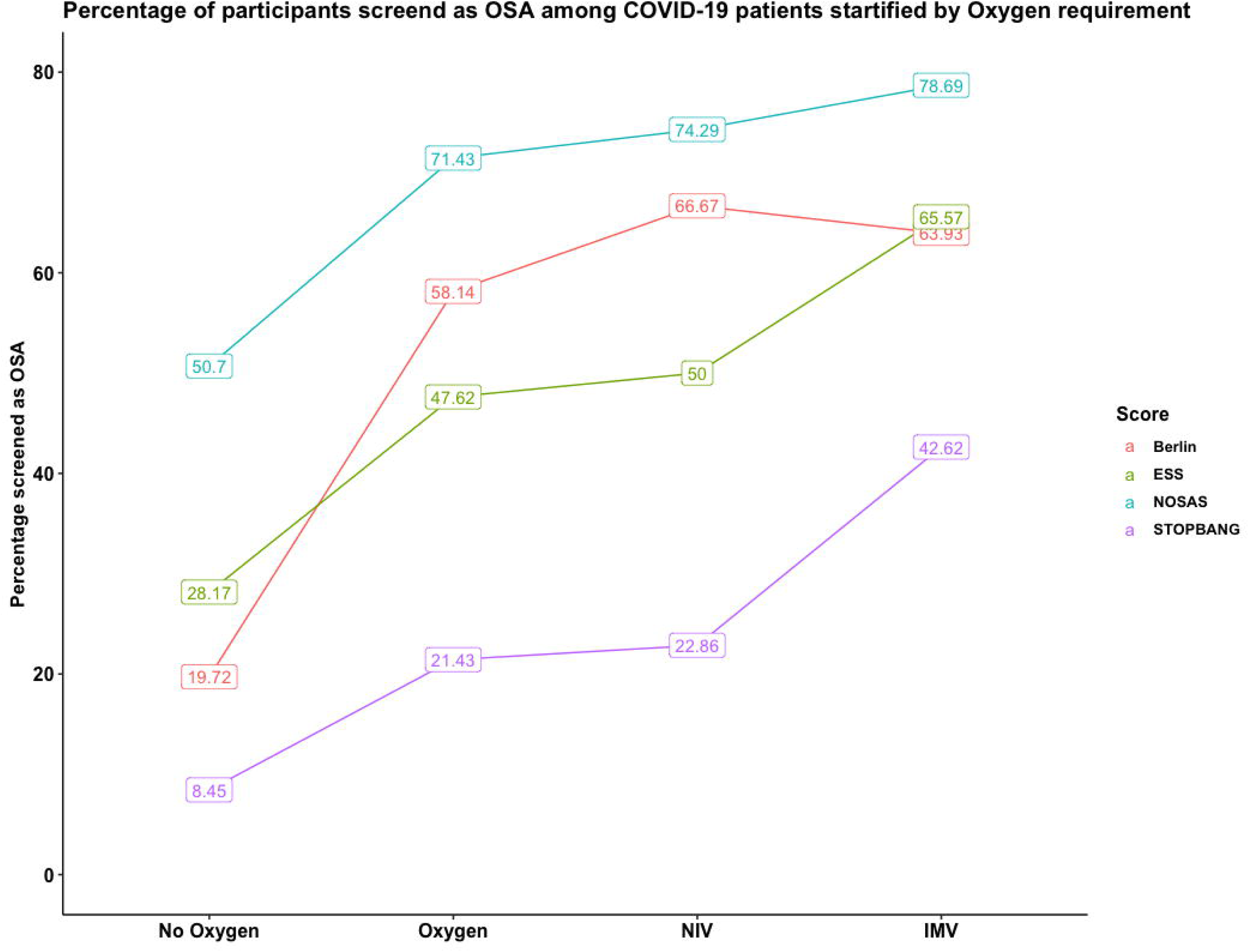
Proportion of patients classified as high risk for OSA by various OSA screening tools across different modes of respiratory support.

On univariate analysis age, Hypertension (HTN), Diabetes Mellitus (DM), Neck circumference and OSA were found to be significant. Since median age was 55 years in our sample, so cut off of 55 years was used for multivariate analysis. On multivariate analysis, STOP BANG was used, since it is most commonly used screening tool for OSA screening both from clinical and research point of view. On binary logistics regression analysis, only age≥55 and STOPBANG score ≥5 were found to be determinants of mortality.

We fitted a logistic model to predict outcome (mortality) with age group, STOP BANG score, presence of diabetes, hypertension, coronary artery disease and neck circumference.The model’s explanatory power is moderate (Tjur’s R^2^ = 0.14). Within this model, the effect of AgeGroup [>=55 years] is positive and can be considered as small and significant (beta = 0.74, SE = 0.37, 95% CI [0.04, 1.48]), while the effect of STOP BANG score in Highriskis also positive and can be considered as small and significant (beta = 0.91, SE = 0.42, 95% CI [0.08, 1.74]).The effects observed for older age and higher STOP BANG score were adjusted for neck circumference as well as history of comorbidities. Odds ratio for these variables which are exponentiated coefficients of logistic model are also presented. Odds of mortality were 2.10 (1.04-4.37, p=0.042) among age more than 55 years compared to those with age less than 55 years. Participants classified in high risk category on STOP BANG score were having odds of 2.48 (1.09-5.69, p=0.031) for mortality compared to those classified as low to intermediate risk for OSA. (Table 3).

**Table 3:** Results of binary logistics regression analysis for determinants of mortality

| Dependent:<br>Deceased |  | Survived | Deceased | OR (univariable) | OR (multivariable) |
| --- | --- | --- | --- | --- | --- |
| AgeGroup | <55 yrs | 84 (84.0) | 16 (16.0) | - | - |
|  | >=55 yrs | 72 (63.7) | 41 (36.3) | 2.99 (1.57-5.90,<br>p=0.001) | 2.10 (1.04-4.37,<br>p=0.042) |
| Gender | Male | 105 (72.9) | 39 (27.1) | - | - |
|  | Female | 51 (73.9) | 18 (26.1) | 0.95 (0.49-1.80,<br>p=0.878) | - |
| BMI | Mean (SD) | 26.6 (4.8) | 27.5 (4.9) | 1.04 (0.98-1.11,<br>p=0.214) | - |
| Neck<br>Circumf | Mean (SD) | 39.6 (5.1) | 41.0 (3.0) | 1.07 (1.00-1.16,<br>p=0.092) | 1.04 (0.97-1.13,<br>p=0.272) |
| STOPBANG | Low to Int. | 129 (80.6) | 31 (19.4) | - | - |
|  | High | 24 (49.0) | 25 (51.0) | 4.33 (2.20-8.66,<br>p<0.001) | 2.48 (1.09-5.69,<br>p=0.031) |
| DM | No | 97 (80.8) | 23 (19.2) | - | - |
|  | Yes | 59 (63.4) | 34 (36.6) | 2.43 (1.31-4.56,<br>p=0.005) | 1.61 (0.78-3.31,<br>p=0.192) |
| HTN | No | 98 (82.4) | 21 (17.6) | - | - |
|  | Yes | 58 (61.7) | 36 (38.3) | 2.90 (1.56-5.50,<br>p=0.001) | 1.27 (0.56-2.86,<br>p=0.567) |
| CAD | No | 142 (75.5) | 46 (24.5) | - | - |
|  | Yes | 14 (56.0) | 11 (44.0) | 2.43 (1.01-5.71,<br>p=0.043) | 1.60 (0.59-4.28,<br>p=0.351) |

Number in data frame = 213, Number in model = 209, Missing = 4, AIC = 228.7, C-statistic = 0.738, H&L = Chi-sq(8) 11.21 (p=0.190)

## Discussion

This is one of the first prospective studies of sequentially hospitalized patients with confirmed COVID 19 status who were screened for possible OSA in a questionnaire-based format. This study shows that OSA could be an independent risk factor for poor outcome in patients with COVID19.

Studies have highlighted association of severity of COVID 19 with older age, obesity, male sex and co-morbidities like DM, HTN, Coronary Artery Disease and Chronic Kidney Disease. Few researchers have observed a possible association with obstructive sleep apnea retrospectively. Brian E. Cade et al analyzed electronic health record data and observed 443 of 4668 participants with sleep apnea had increased mortality rate of 11.7% as compared to controls (6.9%) with an odds ratio of 1.79^23^. In a study involving 700 patients, 124 had pre diagnosed OSA; of all the patients requiring ICU care, 29% patients had pre-existent OSA in the same study^12^. Two small case series focusing critically ill patients of COVID 19 had shown 20-25% patients having OSA^13,14^. In the CORONADO study, 144/ 1189 patients were already known case of OSA. They have also found OSA to be independent risk factor for poor outcome in COVID 19 related illness^24^.

Age and Neck circumference are integral component of most OSA screening questionnaire (STOPBANG, NoSAS and BQ) and presence of hypertension is included as question in STOPBANG and BQ. Thus, multivariate analysis was done to find independent association of OSA with mortality. In-fact in our study, only age and OSA were found to be significant factor for mortality; HTN & DM were not associated with mortality. This was in contrast to most of previous studies, in which HTN & DM were found to be important factors for mortality.

Cardiac morbidity in COVID patients seems to be high and could prove fatal. Cardiac complications in SARS-CoV 2 infection includes myocarditis, cardiomyopathy, acute myocardial infarction, heart failure, venous thromboembolism and arrythmias^25^. These complications might get accentuated in presence of OSA which is a known risk factor for heart failure, acute cardiovascular events, arrhythmias and hypertension. Apart from myocarditis, cardiac arrhythmias remain cause of poor outcome in patients of COVID-19. Atrial Fibrillation and Non-sustained ventricular tachycardia (NSVT) was associated with ICU admission after multivariate adjustment^26^. Hemodynamic alterations in OSA lead to polycythemia and sluggish blood flow, which can possibly lead to procoagulant state^27^. Presence of procoagulant state is a breeding ground for COVID related coagulopathy. OSA is known to be associated with dyslipidemia i.e. increased triglyceride, cholesterol and LDL levels along with reduction in HDL levels^28^. OSA has been associated with obesity, HTN, DM, CAD, arrythmia, chronic kidney disease, dyslipidemia, metabolic syndrome, and pulmonary embolism^29^. All these diseases were almost consistently associated with poor prognosis in patients with COVID19 in various studies^30, 31^.

STOPBANG, BQ and NoSAS are easy to use screening tools for OSA diagnosis and have been shown to have decent sensitivity and specificity compared to Polysomnography (PSG)^32,15,33^. In our study, it was shown that patients with higher respiratory requirements had significantly higher probability of having OSA and this was consistently seen in all questionnaires for increasing severity of respiratory support.

Strength of this study is that screening tools wereasked to both patients and sleeping partner. It is known fact that questions like history of apnea and history of snoring are more reliably answered by patients sleeping partner rather than patients. So, if there was any discrepancy in the answers for apnea or snoring, then answer from sleeping partner was considered final. Those patients who were not in a state to answer questions were excluded from our study. Secondly, we screened for OSA using multiple questionnaires. Most importantly, this is the first study in which patients being admitted for COVID19 were screened for OSA by different questionnaires. All previous studies on possible association between OSA and COVID19 ARDS were done in already diagnosed OSA cases.

Important limitations were:1) It was a single center study conducted in a tertiary care hospital where relatively more sick patients were admitted. This poses possibility of selection bias and therefore results of the study should be interpreted in this context and not be generalized to all COVID-19 patients.2) Our diagnosis of OSA was based on screening questionnaires and gold standardPSG could not be done for confirmation of OSA. However, we have evaluated risk for OSA by multiple questionnaires and results were coherent. We are currently performing sleep study in COVID19 ARDS survivors one month afterdischargeand this will shed more light on the strength of association between COVID19 ARDS and OSA.

## Conclusions

This study shows that OSA might be an independent risk factor for poor outcome in COVID 19 related illness.

## Data Availability

The data referred in study is available and can be provided if required.

## Acknowledgements

Dr. Abhishek Goyal MD, DM had full access of the data and takes responsibility for the integrity of the data and accuracy of data analysis. He is the guarantor of the content of the manuscript, including the data and analysis.

Dr. Avishek Kar and Dr. Khushboo Saxena contributed substantially and equally to the study design, data analysis and interpretation, and the writing of the manuscript.

## Contributions of authors

Goyal Abhishek: conceptualization, revision of final manuscript for intellectual data, statistical analysis and interpretation

Kar Avishek: Data collection, managed patients, drafted manuscript

Saxena Khushboo: Managed patients, data collection, drafted manuscript

Pakhare. Abhijit: Statistical analysis and interpretation of data

Khurana Alkesh: Managed patients, data collection

Saigal Saurabh: Managed patients, data collection

